# Optimal vaccine allocation for COVID-19 in the Netherlands: a data-driven prioritization

**DOI:** 10.1101/2021.07.20.21260889

**Authors:** Fuminari Miura, Ka Yin Leung, Don Klinkenberg, Kylie E. C. Ainslie, Jacco Wallinga

**Affiliations:** Centre for Infectious Disease Control, National Institute for Public Health and the Environment (RIVM), Bilthoven, the Netherlands; Center for Marine Environmental Studies (CMES), Ehime University, Ehime, Japan; School of Public Health, Imperial College London, London, United Kingdom; MRC Centre for Global Infectious Disease Analysis and Abdul Latif Jameel Institute for Disease and Emergency Analytics, Imperial College London, London, United Kingdom; Department of Biomedical Data Sciences, Leiden University Medical Center (LUMC), Leiden, the Netherlands

**Author notes:** **Author contributions** Conceptualization: FM JW. Data curation: FM. Formal analysis: FM KL JW. Investigation: FM. Methodology: FM KL DK JW. Software: FM. Validation: FM KL JW. Visualization:FM. Writing – original draft: FM KL DK KA JW. Writing – review & editing: FM KL DK KA JW. **Data Availability** All data are available from GitHub repository (https://github.com/fmiura/VacAllo_2021). **Funding** FM acknowledges funding from Japan Society for the Promotion of Science (JSPS KAKENHI, Grant Number 20J00793). This project has received funding from the European Union’s Horizon 2020 research and innovation programme - project EpiPose (grant agreement number 101003688). **Competing interests** The authors have declared that no competing interests exist.

## Abstract

For the control of COVID-19, vaccination programmes provide a long-term solution. The amount of available vaccines is often limited, and thus it is crucial to determine the allocation strategy. While mathematical modelling approaches have been used to find an optimal distribution of vaccines, there is an excessively large number of possible schemes to be simulated.

Here, we propose an algorithm to find a near-optimal allocation scheme given an intervention objective such as minimization of new infections, hospitalizations, or deaths, where multiple vaccines are available. The proposed principle for allocating vaccines is to target subgroups with the largest reduction in the outcome of interest, such as new infections, due to vaccination that fully immunizes a single individual. We express the expected impact of vaccinating each subgroup in terms of the observed incidence of infection and force of infection. The proposed approach is firstly evaluated with a simulated epidemic and then applied to the epidemiological data on COVID-19 in the Netherlands.

Our results reveal how the optimal allocation depends on the objective of infection control. In the case of COVID-19, if we wish to minimize deaths, the optimal allocation strategy is not efficient for minimizing other outcomes, such as infections. In simulated epidemics, an allocation strategy optimized for an outcome outperforms other strategies such as the allocation from young to old, from old to young, and at random. Our simulations clarify that the current policy in the Netherlands (i.e., allocation from old to young) was concordant with the allocation scheme that minimizes deaths.

The proposed method provides an optimal allocation scheme, given routine surveillance data that reflect ongoing transmissions. The principle of allocation is useful for providing plausible simulation scenarios for complex models, which give a more robust basis to determine intervention strategies.

**Author summary:** Vaccination is the key to controlling the ongoing COVID-19 pandemic. In the early stages of an epidemic, there is shortage of vaccine stocks. Here, we propose an algorithm that computes an optimal vaccine distribution among groups for each intervention objective (e.g., minimizing new infections, hospitalizations, or deaths). Unlike existing approaches that use detailed information on at-risk contacts between and among groups, the proposed algorithm requires only routine surveillance data on the number of cases. This method is applicable even when multiple vaccines are available. Simulation results show that the allocation scheme optimized by our algorithm performed the best compared with other strategies such as allocating vaccines at random and in the order of age. Our results also reveal that an allocation scheme optimized for one specific objective is not necessarily efficient for another, indicating the importance of the decision-making at the early phase of distributions.

## Introduction

SARS-CoV-2 has posed a great threat to public health. As of 8 July 2021, 33,270,049 cases and 740,809 deaths with COVID-19 have been reported in the EU/EEA (1), and globally there have been 4,006,882 deaths reported (2). While non-pharmaceutical interventions (NPIs) reduce transmission (3,4), the societal cost of implementing these measures is enormous (5,6), and the effect is short-lived. Vaccination offers a long-term approach to control COVID-19.

Currently, sixteen vaccines have been approved for use, 99 companies are still conducting clinical trials to develop next generation vaccines (7). There is a limited amount of vaccine available, especially in low- and middle-income countries, because of narrow production capacity and logistics (2,8,9). There is an urgent need to optimize the allocation of scarce vaccines.

The optimal allocation depends on the objective of infection control. If the objective is to minimize hospitalizations, it might be best to target those with the highest risk of severe illness upon infection. If the objective is to reduce transmission of infection, it might be best to target the individuals who contribute most to future infections. Similar allocation problems were previously explored for influenza vaccination programmes (10–12). The allocation of COVID-19 vaccines has been evaluated in combination with NPIs (13–15), with age-varying vaccine efficacy (16), and with different sizes of the vaccine stockpile (17,18). These studies examined plausible scenarios with numerous combinations of models and parameters; however, the challenge here is that there are innumerable possible allocation schemes to compare.

Here we show a data-driven approach to find optimal allocation schemes, by age group and vaccine type, that minimize either new infections, hospitalizations, or deaths. As per previous studies (13–18), we stratify the population by age, because age is shown to be an important risk factor for susceptibility (19,20), severe illness (21,22), and mortality (21,23,24). We apply the proposed approach to a simulated epidemic to evaluate its performance. We also test it with epidemiological data of COVID-19 in the Netherlands, in order to find optimal allocation schemes for different types of vaccines.

## Results

### Impact of a single unit of vaccination

We are interested in prioritizing a subgroup, to target vaccination of individuals in group *i*, by considering within- and between-subgroup transmissions. To find optimal allocation schemes, the proposed approach relies on establishing the impact of a single unit of vaccine (i.e., the number of doses to fully immunize a single individual), as described in the following three steps.

First, we write an age-stratified transmission process in matrix form by introducing the next generation matrix ***K*** (25–27). The next generation matrix ***K*** gives the number of new infections in a successive generation, such that the number of new infections at time *t* + 1 after one generation of infections is ***x***(*t* + 1) = ***Kx***(*t*). Note that ***K*** is a *m* × *m* matrix, and we have *m* age groups. We start with a *m* × 1 vector of age-specific infection at time *t*, ***x***(*t*).

Second, we define the “impact” of a single unit of vaccination as the reduction in the number of new infections generated by an infected individual. A decrease in the number of infected individuals at time *t* + 1, ***x***(*t* + 1), is expressed as a result of changes in the next generation matrix ***K*** and in the number of infected individuals ***x***(*t*) due to vaccinating one individual. With simplified notation, we can write this as ***x***′(*t* + 1) = ***K***′***x***(*t*) + ***Kx***′ (*t*), where ***K***′ and ***x***′ (*t*) are derivatives with respect to the number of vaccinated individuals; ***K***′***x***(*t*) is the direct effect of vaccinating an individual and removing them from the susceptible population and ***Kx***′ (*t*) is the indirect effect of vaccinating a single individual by reducing onward infections (see Eq.S4 and Eq.S7 for full notation).

Third, the main interest here is to approximate the next generation matrix ***K*** using observed epidemiological data. By approximating ***K***, we can calculate above-defined changes without knowing the detailed contact information between groups. To derive the approximated form, we require that at-risk contacts are reciprocal. With this condition, the next generation matrix ***K*** can be safely approximated by the combination of the force of infection 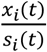 (i.e., incidence rate of new infections *x*_*i*_(*t*) per susceptible individual *s*_*i*_(*t*)) and the incidence rate of new infections per individual 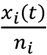, and its approximation error is guaranteed to be small if the observation interval for new infections is more than two generation intervals (28) (see detailed derivation in **SI Text-1**).

Using the above results, when age group *i* is targeted for vaccination, its impact can be measured as the contribution of the change in group *i* to the relative reduction in the number of new infections after one generation of infection (see Eq. S11 in **SI Text-1**). As a result, we can define this quantity as the “importance weight” of infection 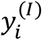, given by

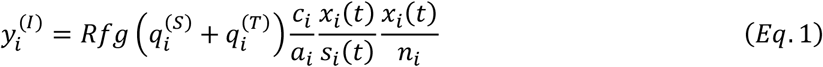

where *R* is the reproduction number, *f* and *g* are normalizing constants, 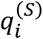 and 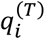 are vaccine efficacies for susceptibility and transmissibility in age group *i, c*_*i*_ is per contact probability of transmitting infection for age group *i*, and *a*_*i*_ is per contact probability of acquiring infection for age group *i*. We can interpret the quantity 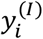 as the expected reduction in the number of new infections generated by an infected individual after introducing a single unit of vaccine in group *i*, compared with the counterfactual situation where no vaccine is introduced.

The importance weight can be generalized for other disease outcomes. We find that the generalized form of *Eq*.1 for other disease outcomes can be written as the product of the relative change in the number of new infections 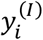 and a disease progression rate (see the derivation in **SI Text-1**). To illustrate its application, we introduce the importance weight of hospitalization 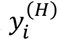 and death 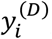, which are defined as the relative reduction in the number of hospitalizations and deaths;

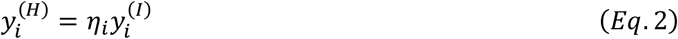

and

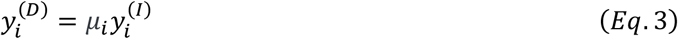

where *η*_*i*_ is the infection hospitalization rate and *μ*_*i*_ is the infection mortality rate for age group *i*.

### Prioritization algorithm

Given a limited stockpile of vaccines, we assess the expected impacts of a single vaccination on the number of new infections, hospitalization, or deaths, with importance weights (i.e., 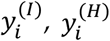 and 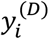 shown in ***Eq***.1-3). In the case that there are multiple types of vaccines, we can define importance weights by vaccine type. To illustrate the algorithm proposed in this study, we use the example of minimization of hospitalization, letting 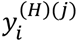 denote the importance weight of hospitalization (H) for vaccine type *j* in age group *i*. By comparing age and vaccine type specific importance weights, the sequential allocation is performed as described below:

Step-1: Decide the objective of infection control (in this example, minimizing hospitalization (H))

Step-2: Calculate importance weights 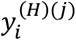 per age-group *i* and vaccine type *j*

Step-3: Find a combination of age-group *i* and vaccine type *j* that has the largest importance weight; this provides the selected age group and selected vaccine type.

Step-4: Allocate a single unit of the selected vaccine to the selected age-group

Step-5: Re-calculate importance weights by decreasing the weights in the targeted age-group, as 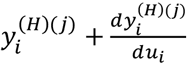. Others remain the same.

Step-6: Repeat above until the end of vaccine stockpile.

Note that in step-5 all the importance weights of the age group *i* are updated. This is because the allocation of one vaccine type depletes susceptible and infectious individuals in the targeted age group, and thus it affects the expected impacts of other vaccines from next iterations (see detailed derivation in **SI Text-1**). The pseudo code for this algorithm is provided in **Table-S2**.

There are four conditions that should be met; (i) the epidemic grows exponentially over the time interval, (ii) at-risk contacts are reciprocal, (iii) the observation interval for new infections is sufficiently long, and (iv) there is no major change in the age distribution of the risk of infection. With these assumptions, we can reconstruct the (approximated) next generation matrix and calculate the expected impact on each outcome due to vaccination, without detailed information about contacts between groups (28).

### Test against simulated data

We test the performance of the proposed algorithm using a simulated epidemic. **Figure-1(A)** illustrates the generated epidemic curve where we set the basic reproduction number *R*_0_ to 1.2 and the generation time as 5 days, based on the estimates of SARS-CoV-2 infections, following previous modelling studies (13,16) (see **Method** for details of simulation settings). Although only partial observations on the incidence and force of infection are used as inputs, the allocation strategies yielded by our algorithm perform better than other strategies that we tested in most cases (i.e., random allocation, allocation from young to old groups, allocation from old to young groups, and no vaccination) (**Figure-1(D)-(F)**).

**Fig-1.**
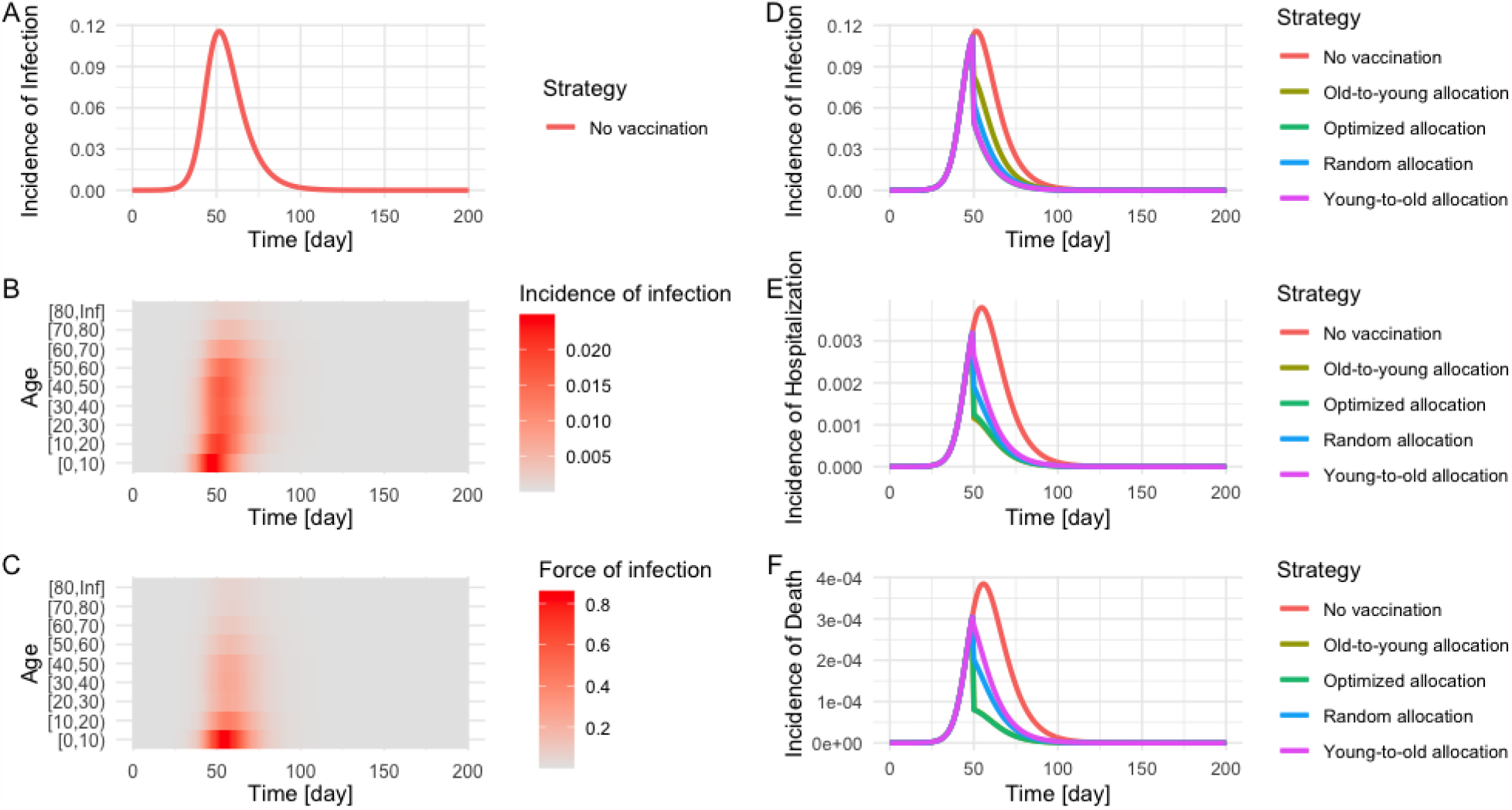
Simulated epidemic and evaluation of the impact of vaccination by allocation strategy. The epidemic is simulated by an age-structured SIR model. *R*_0_ and generation time were set as 1.2 and 5 days, respectively. The population was stratified by 10-year age bin, and a contact matrix of the Netherlands in June 2020 was used for the simulation (32). Panel **(A)** illustrates the total incidence of infection in the population, and age-specific incidences **(B)** and the force of infection **(C)** reflect heterogeneous contacts between age-groups. The impact of vaccination on the number of infections **(D)**, hospitalizations **(E)**, and deaths **(F)** was compared under five different strategies; no vaccination (red), allocation from old to young groups (yellow), young to old groups (purple), at random (blue), and optimized allocation (green). For simplicity, the vaccination coverage was set as 40%, and the effect of vaccines was in place at day 50 (from the initial time point of the simulation), resulting in the immediate depletion of susceptible and infected individuals on that day.

### Age distribution of allocated vaccines by prioritization scheme

We apply the proposed approach to epidemiological data on COVID-19 in the Netherlands. Higher efficacious vaccines are allocated first, and then lower efficacious vaccines are distributed later on (**Figure 2** and **Figure S2**). **Figure 2** shows the detailed breakdown of allocated vaccines by age group and vaccine type in each allocation scheme, and all the schemes start with the highest efficacious vaccine (i.e., Pfizer vaccine). Since high vaccine efficacy results in larger impacts per vaccination (**Eq-2**), it is natural to prioritize the allocation of higher efficacious vaccines.

**Fig-2.**
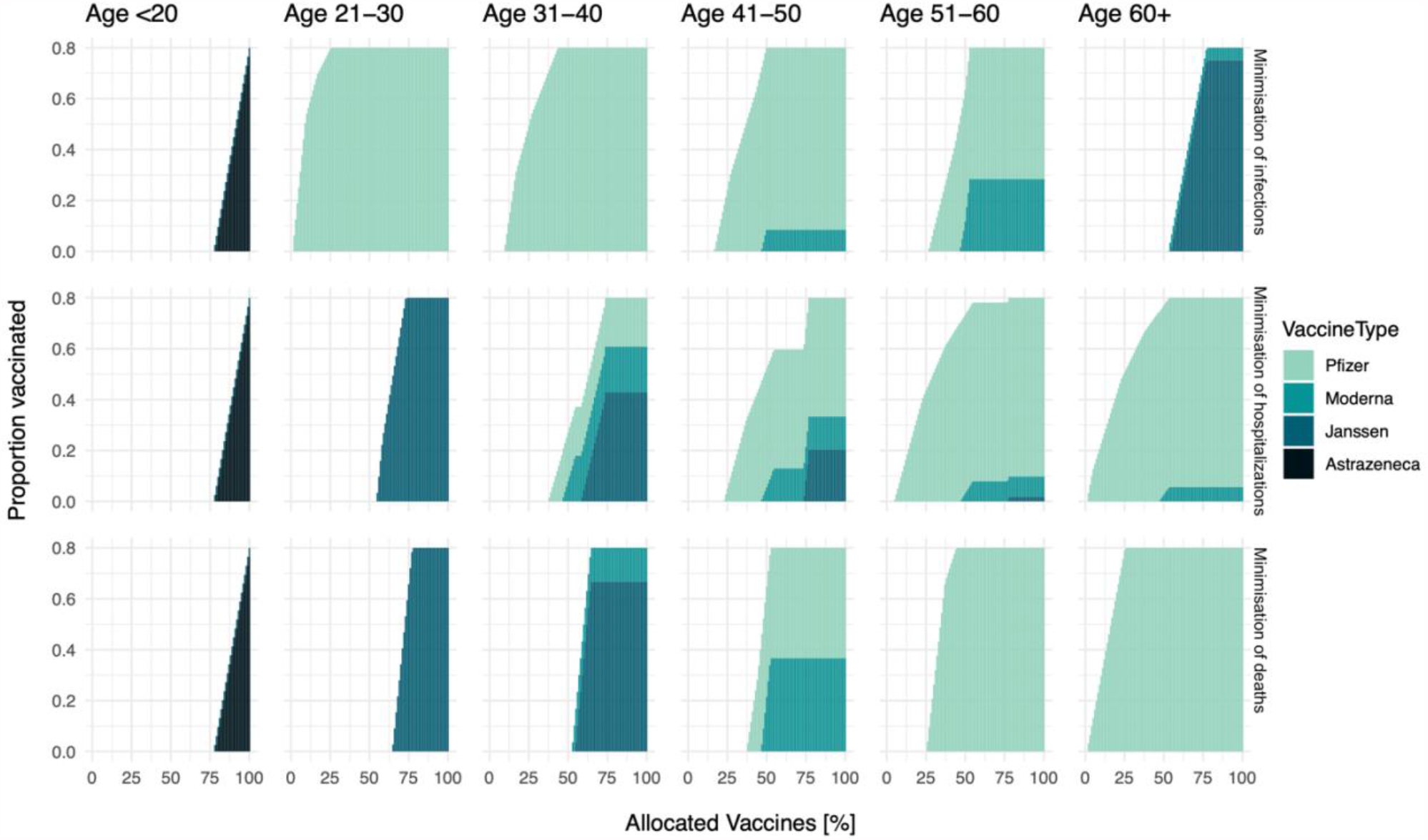
The order of vaccine allocation by age and by prioritization strategy for a stockpile that suffices to vaccinate 80% of the population. From the top row, the objective is the minimization of infections, hospitalizations, and deaths respectively. From the left column, the proportion of vaccinated among age <20, 21-30, 31-40, 41-50, 51-60, 60+ are plotted over allocated vaccines. Note that the X-axis shows the percentage of allocated vaccines.

Depending on the objective of infection control, the type of vaccines that each age group receives would differ. If a specific age group is significantly contributing to the objective, it is better to distribute higher efficacious vaccines to that group. For example, there is a large contribution of age 21-30 for the number of infections (**Figure S1**), and thus higher efficacious vaccines are distributed to that group if the objective is to minimize the number of infections (top row in **Figure 2**). If we wish to minimize the number of hospitalizations or deaths, those vaccines would be distributed to the elderly (second and third rows in **Figure 2**).

The optimal timing of switching from one age group to another also varies by objective. When we set the objective as the minimization of the number of infections or hospitalizations, the selected allocation orders for these two objectives suggest to distribute vaccines to several age-groups in parallel (first and second rows in **Figure 2** and **Figure S3**). By contrast, when we set the objective as the minimization of the number of deaths, the allocation scheme generally focused on one age group, from old to young, and did not switch to the next age group until the vaccination of the first age group (i.e., age 60+) is finished (third row in **Figure 2** and **Figure S3**). In terms of the order and the switching timing, the selected allocation scheme that minimizes deaths is concordant with the current allocation policy in the Netherlands (29).

### Different benefits between vaccine prioritization strategies

Allocation schemes that are optimized for one objective may not be optimal with respect to another, as illustrated by our simulations. If we choose to minimize the number of infections, that allocation scheme is not efficient for the minimization of deaths (**Figure 3 (A)**). In contrast, if we wish to minimize the number of hospitalizations or deaths (**Figure 3 (B)** and **(C)**), those strategies are not efficient for minimizing infections. Especially, the difference in the expected reduction is larger at the early phase of allocations; this is because mainly younger age groups are drivers of transmission (**Figure S1 (A)**), while younger individuals are not in high-risk groups in terms of hospitalization or death (**Figure S1 (F)** and **(G)**).

**Fig-3.**
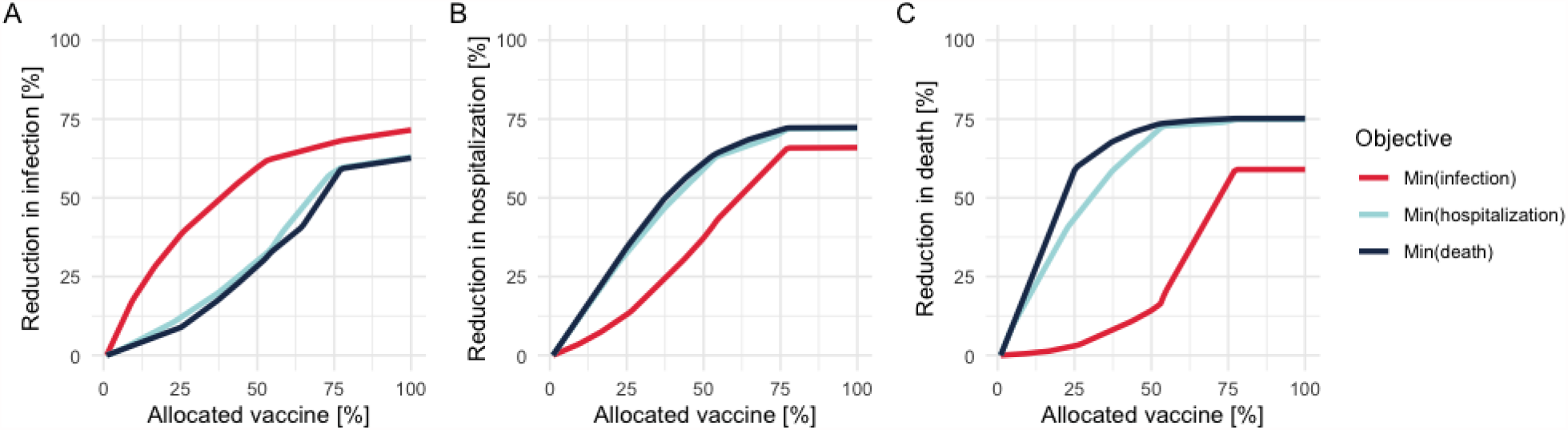
Performance of allocation schemes on different objectives for a stockpile that suffices to vaccinate 80% of the population. The Y-axis shows the percentage reduction in the number of infections (A), hospitalizations (B), and deaths (C), and the X-axis is the percentage of allocated vaccines. Red, light blue, and dark blue plots indicate the allocation strategies to minimize the number of infections, hospitalizations, and deaths respectively. The starting point of effective reproduction number (i.e., the reference point without any vaccination) was set as 1.2.

The proposed algorithm finds the best solution at each allocation step. This results in an optimal solution for small stockpiles, but this local optimal solution is not necessarily optimal for larger stockpiles (so called “greedy algorithm” (30)). To elucidate this property, we simulate an alternative situation, before the approval of the Janssen vaccine, where the breakdown of the stock is Pfizer (40%), AstraZeneca (40%), and Moderna (20%). **Figure S4** illustrates that the allocation scheme to minimize infections results in nearly equal reduction of infections at the end of allocations compared to the other two schemes, although it performed best at the beginning phase.

## Discussion

The present study proposes a prioritization algorithm that can find an optimal allocation of vaccines to different age groups, even with a limited amount of data. Our simulation results show how optimal allocation differs depending on the objective. We apply the algorithm to available Dutch epidemiological data on COVID-19, and the allocation scheme that minimizes deaths is concordant with the current policy in the Netherlands that allocates vaccines from old to young (29).

The proposed method provides first principles to find optimal allocation schemes with limited data, and the output can also be used as a complementary tool to existing computational approaches. Previous studies hinged on dynamic modeling to determine the prioritization of vaccine allocation (13,16,17), and our algorithm can inform a near-optimal distribution of vaccines as input values for those simulations. The proposed method can be used as a cross-check of assumptions in dynamic models, because it does not require the detailed information on contact matrices or non-pharmaceutical interventions. In the COVID-19 pandemic, we have already observed immediate changes of the age distribution of reported cases (20,31), and contact patterns during lockdown are different from usual patterns (32). The strength of our approach is that it relies only on routine surveillance data.

Choosing a different objective for COVID-19 control implies choosing a different optimal allocation scheme. In the case of SARS-CoV-2 infection, individuals who are at higher risk of severe illness and who transmit are different (19,22). Our results (**Fig-1** and **Fig-3**) illustrated that, if we weigh an objective (e.g., minimization of infections) and choose a strategy, the selected scheme is not necessarily efficient for the other objectives (e.g., minimization of hospitalizations and deaths). In our analysis, the difference in the reduction of each outcome was larger at the earlier phase of vaccinations (**Fig-3**), indicating the importance of decision-making in the beginning stage of allocations. While vaccine rollout has progressed rapidly in the first half of 2021 in high-income countries, there is large vaccine inequity globally (33). In many low- and middle-income countries vaccine rollout is hindered by limited supply. An algorithm, such as the one presented here, can be very useful to prioritize vaccine allocation in those countries where maximum impact on disease outcomes must be achieved by a small supply of vaccines. Besides, the proposed method can be easily generalized for a wider range of objectives, by multiplying a disease progression rate (**SI Text-1**). The contribution of this study is to provide a solution how to determine the subgroup with the largest contribution to different outcomes, given limited data.

When the proposed algorithm is applied, several assumptions and underlying conditions of input values should be checked. First, confirmed case counts may not reflect the actual infection dynamics in the population, depending on the level of ascertainment (34,35). Our approach relies on the estimates of group-specific incidence and force of infection, as the best proxy of ongoing transmission, and thus potential biases in the surveillance should be carefully scrutinized. Second, our modelling simplified offering vaccine doses as a single event and parameterized vaccine efficacies as the ability of reducing infections (***Q***_***S***_) and blocking transmissions (***Q***_***T***_), separately. While there is an advantage to be able to evaluate various characteristics of vaccines by incorporating both the marginal benefit and direct protection, additional supportive evidence on the vaccine efficacy is required. Third, we assume that risk contacts are reciprocal and that individuals are randomly mixing in each group. Although the reciprocity is not violated by a broad class of diseases (32,36), if there were a specific age group that refuses vaccinations, and if its proportion became significantly large, that kind of clustering effect might influence the result of approximation of transmission processes.

In conclusion, the present study proposes an approach to find an optimal allocation of vaccines for various objectives, given routine surveillance data. The principle of allocation is simple and interpretable. These features are essential for decision making and for answering to ethical questions that are inherent to allocation of scarce resources. In the context of COVID-19 control, the ability to base important decisions on real-time data, rather than the assumed effect of contact patterns and non-pharmaceutical interventions, might provide a more robust scientific basis for COVID-19 control.

## Data Availability

All data are available from GitHub repository (https://github.com/fmiura/VacAllo_2021).

## Acknowledgement

We thank Jantien Backer for sharing the data on contact matrices in the Netherlands, and Scott McDonald for helpful discussions.

## Materials and Methods

### Covid-19 epidemic data in the Netherlands

The population data was stratified into six age groups [<20, 21-30, 31-40, 41-50, 51-60, 60+]. For each age group, we used data on the population size, seroprevalence, incidence of notified cases, maximum vaccine uptake (i.e., willingness to be vaccinated), COVID-19 hospitalization rate, COVID-19 mortality rate, and vaccine efficacy against infection and transmission (**Figure S1**). The seroprevalence data was obtained from the Pienter-Corona study among a representative sample of the Dutch population, collected in June 2020 (37). We used this data to calculate the proportion of susceptible individuals per group, that is, 1 – seropositive rate. We used infection hospitalization rate and infection mortality rate that were estimated by published studies based on pooled analyses over 45 countries (22,24) rather than specific estimates for the Netherlands.

The maximum vaccine uptake was assumed to be 80% for all age groups. The vaccine efficacy was assumed constant over age-groups (38–41). We assumed the same vaccine efficacies against infection and transmission (**Figure-S1**). To calculate the expected decrease in the number of new infections, hospitalizations, and deaths, as a function of the number of allocated vaccines, the starting point of effective reproduction number *R* (i.e., the reference point without any vaccination) was set to 1.2.

We allocated a vaccine stockpile that covers 80% of the total population. The breakdown of the stock is Pfizer (46%), AstraZeneca (22%), Moderna (8%), and Janssen (24%). Note that we considered the unit of vaccines as a set of full doses; for example, the Pfizer vaccine needs to be administered twice, and the set of those two doses was defined as a single unit here. We assumed that one person can receive only one type of vaccines. Thus, 80% of the population was vaccinated when all vaccines were allocated.

### Performance evaluation with simulated epidemics

We simulated an epidemic, using a deterministic SIR model, where all parameters were known a priori. We evaluated five different allocation strategies: optimal allocation for each objective (i.e., minimization of infections, hospitalizations, and deaths) determined by the proposed algorithm; random allocation; allocation from young to old groups; allocation from old to young groups; and no vaccination. To quantify the impact of vaccinations in each strategy, we took the “no vaccination” scenario as a natural reference point. The population was stratified by 10 year age group, since a contact matrix of the Netherlands in June 2020 was available with those age bins and used for the simulation (32). An age-structured SIR model was used to generate an epidemic curve where *R*_0_ was set as 1.2 with the fixed generation time as 5 days, based on the estimates of SARS-CoV-2 infections following previous modelling studies (13,16). For simplicity, per contact probability of acquiring infection (*a*_*i*_) and per contact of transmitting infection (*c*_*i*_) were assumed to be equal, and the vaccine efficacy was 0.946 based on the estimate for Pfizer (38). The available vaccine stock was set as 40% coverage of the population, which covers a half of the population that are willing to get vaccinated.

As a practical application, observable information (i.e., the incidence of infection and the force of infection) until day 45 was used as inputs, where day 0 is the initial time point of a simulated SIR epidemic. The optimal distribution of vaccines to each age group was yielded by the proposed algorithm. Note that the algorithm does not use the contact matrix. In each scenario, the effect of allocated vaccines became in place at day 50 all at once, resulting in the immediate depletion of susceptible and infected individuals in the population. Replication code is available on GitHub (https://github.com/fmiura/VacAllo_2021).

### Derivation of importance weights

For a broad class of compartmental models, the disease transmission is described as transitions from discrete states (e.g., susceptible-infectious-recovered states in the SIR model), and the dynamics is generated by a system of nonlinear ordinary differential equations (ODEs) that depicts the change over time. By linearizing ODEs, any (linear) system can be described by a matrix form (26). Within this linearized subsystem, one can determine the reproduction number *R* as the dominant eigenvalue of the next generation matrix ***K*** (25–27).

The first step is to relate the observed data to the next generation matrix ***K***. If at-risk contacts are reciprocal, the next generation matrix ***K*** becomes a product of symmetric matrices and diagonalizable. This condition allows the decomposition of ***K***, and thus we can approximate ***K*** by the top left and right eigenvectors that can be (approximately) described by the incidence of new infections and force of infection (28).

Once the matrices are specified, we can evaluate the impact of a single unit of vaccination, as the sensitivity (or elasticity) of the transition matrix (see the general idea of the sensitivity of a matrix in (42), and its application in infectious disease epidemiology in (27,43)). The change in the number of infections per single vaccination can be formulated as the result of depletion of susceptible and infectious individuals from the population (**Eq-S4** in **Text-S1**), and subsequently, we obtain its effect on the dominant eigenvalue of the next generation matrix that was already introduced in the first step as an approximation with observed data. The expected impact here is defined as the importance weight; if we allocate a single unit of vaccine to the group with the largest importance weight, that results in the minimization of the dominant eigenvalue, that is, the expected number of infections, hospitalizations, or deaths in total.

## Supporting information

**Figure S1**. Age-specific input data

**Figure S2**. Simulated vaccine allocations by age and by vaccine type

**Figure S3**. Simulated prioritization of age-group by allocation scheme

**Figure S4**. Simulated impact of vaccinations

**Text S1.** Mathematical details

**Figure S1.**
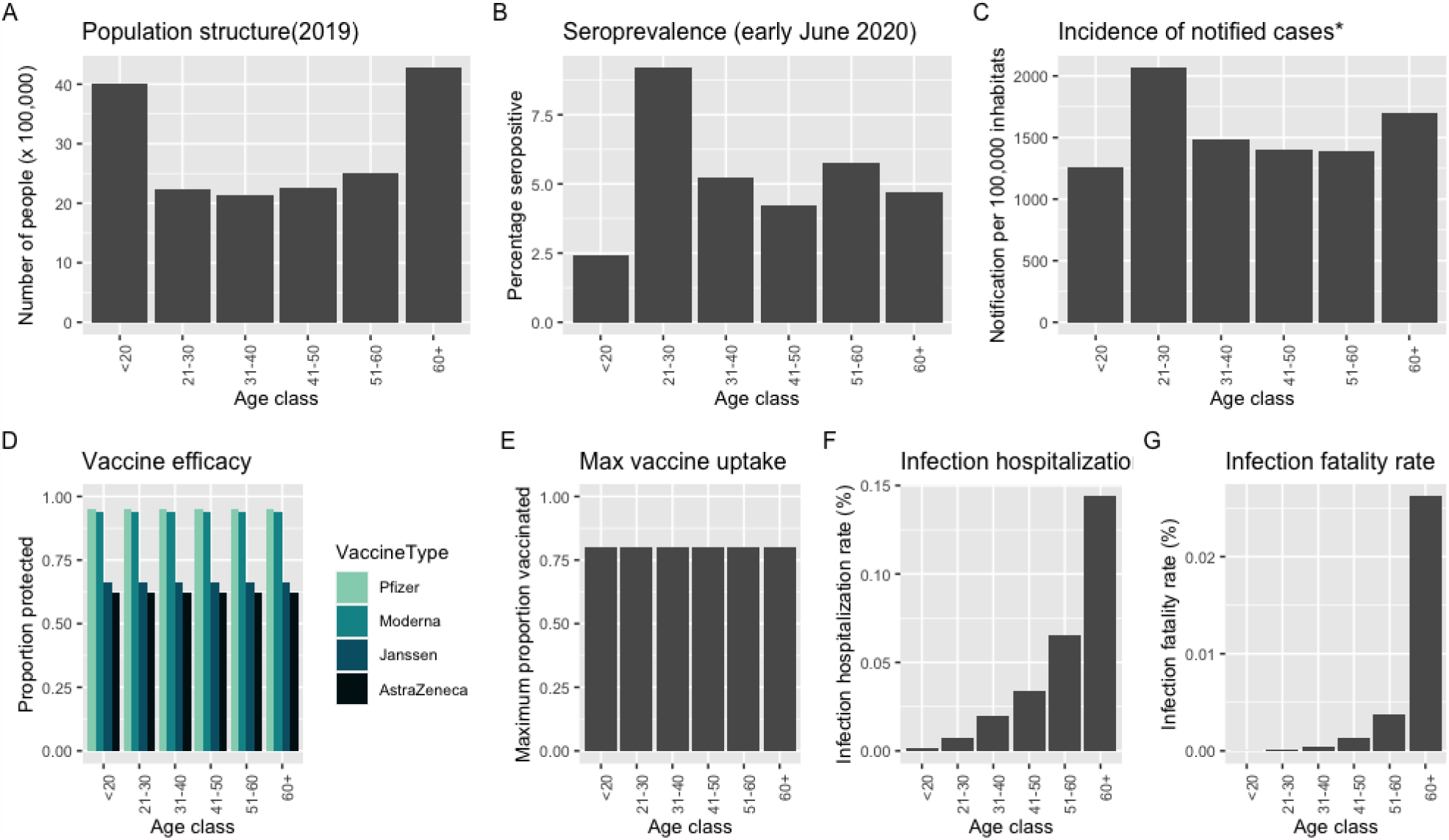
Age-specific input data for the proposed algorithm to obtain optimal allocation schemes. (A) Population structure in the Netherlands in 2019 (B) Seroprevalence observed in the Pienter-Corona study among a representative sample of the Dutch population in June (37). (C) Incidence of notified cases, in 30 days before October 19, 2020 (D) Vaccine Efficacy by vaccine type. From lighter to darker blue, bars indicate Pfizer Moderna, Janssen, AstraZeneca. Note that the constant efficacy by age here is an assumption, based on reported over all vaccine efficacies (38– 41). (E) Maximum vaccine uptake per age group. 80% for all groups is assumed here. (F) COVID-19 hospitalization rate. These values are based on (22). (G) COVID-19 mortality rate. These values are based on (24).

**Figure S2.**
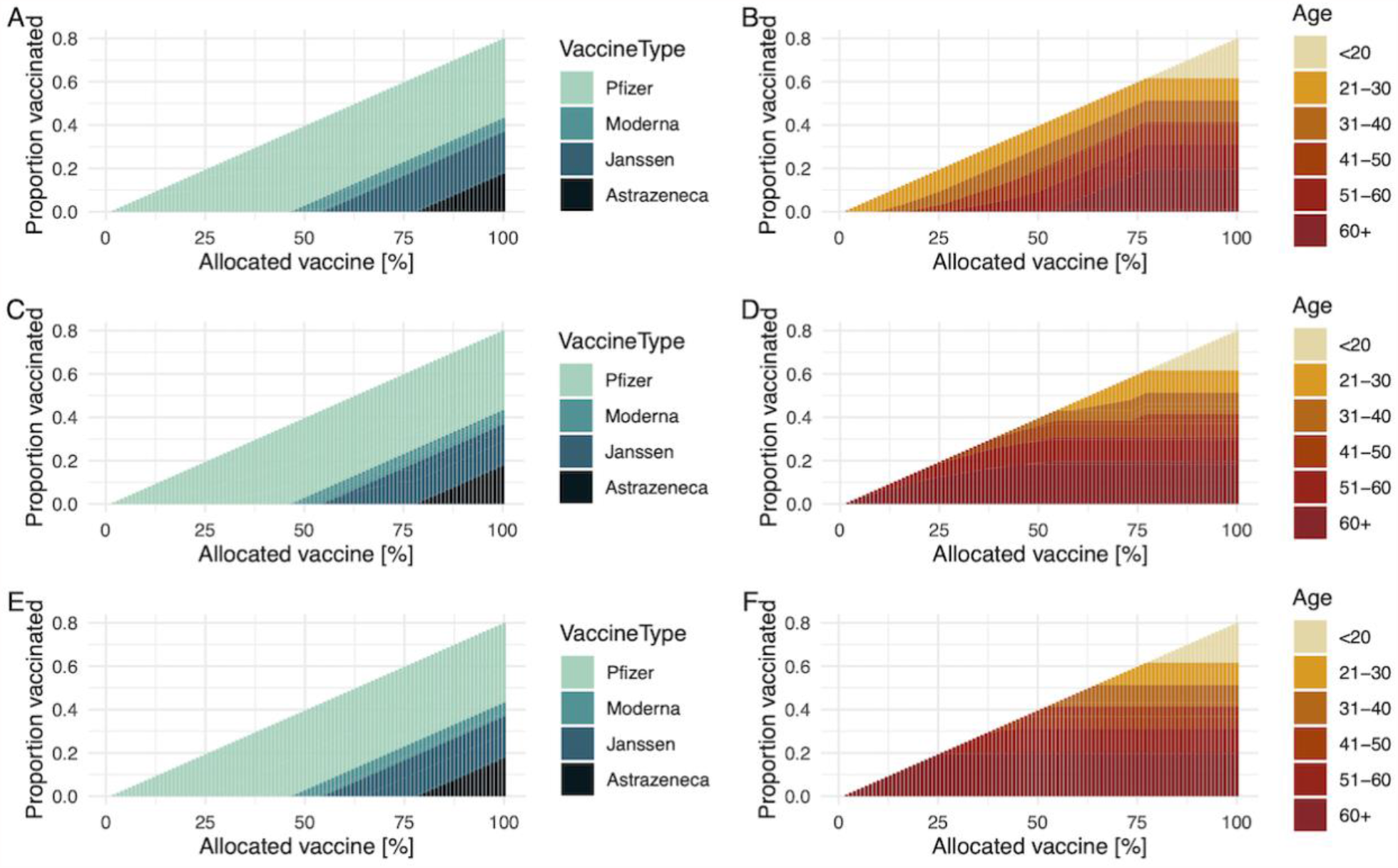
Vaccine allocations based on simulated data when the objective is to minimize the number of infections ((A) and (B)), hospitalizations ((C) and (D)), and deaths ((E) and (F)). In left three panels, from lighter to darker blue, bars indicate Pfizer Moderna, Janssen, AstraZeneca. In right three panels, the darker color shows the older age groups, and age bins are [20<,21-30,31-40,41-50,51-60,60+].

**Figure S3.**
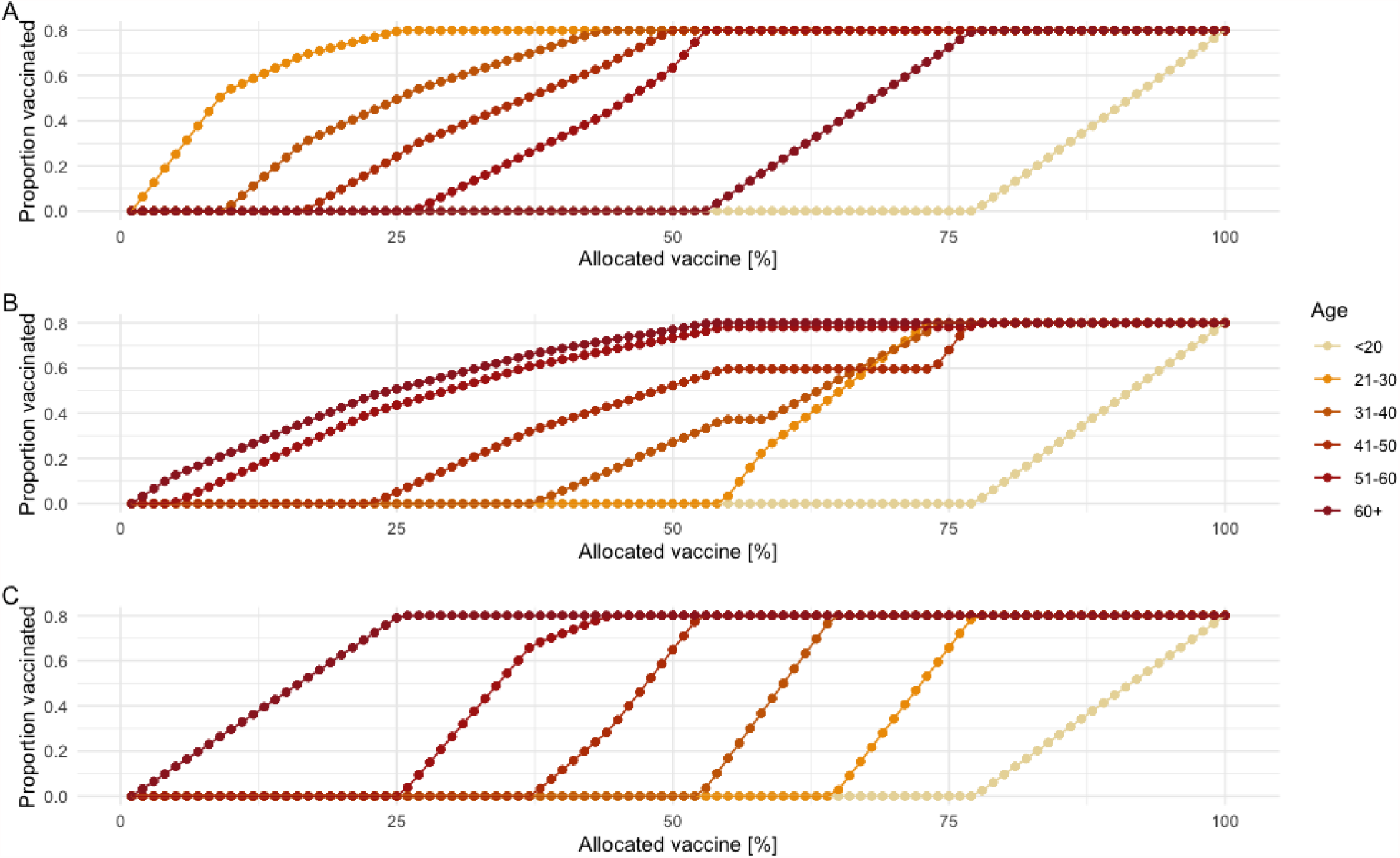
Vaccine allocation based on simulated data when the objective is to minimize the number of infections (A), hospitalizations (B), and deaths (C). The darker color shows the older age groups, and age bins are [20<,21-30,31-40,41-50,51-60,60+].

**Figure S4.**
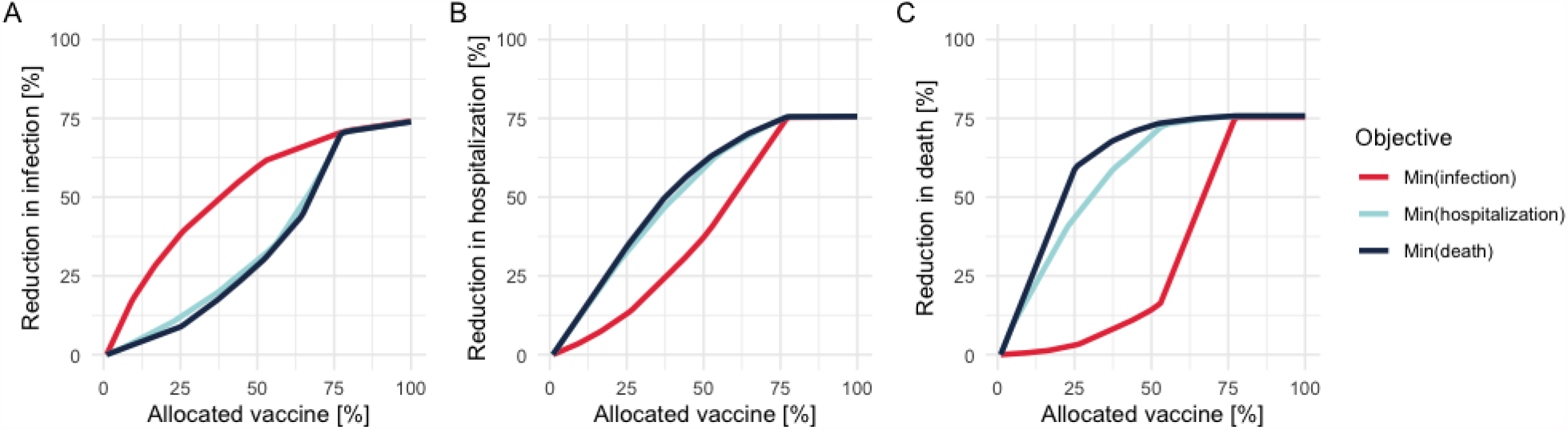
Performance of allocation schemes on different objectives for a stockpile that suffices to vaccinate 80% of the population. The breakdown of the stock is Pfizer (40%), AstraZeneca (40%), and Moderna (20%). The Y-axis shows the percentage reduction in the number of infections (A), hospitalizations (B), and deaths (C), and the X-axis is the percentage of allocated vaccines. Red, light blue, and dark blue plots indicate the allocation strategies to minimize the number of infections, hospitalizations, and deaths respectively. The starting point of effective reproduction number (i.e., the reference point without any vaccination) was set as 1.2.

**Table S1.**
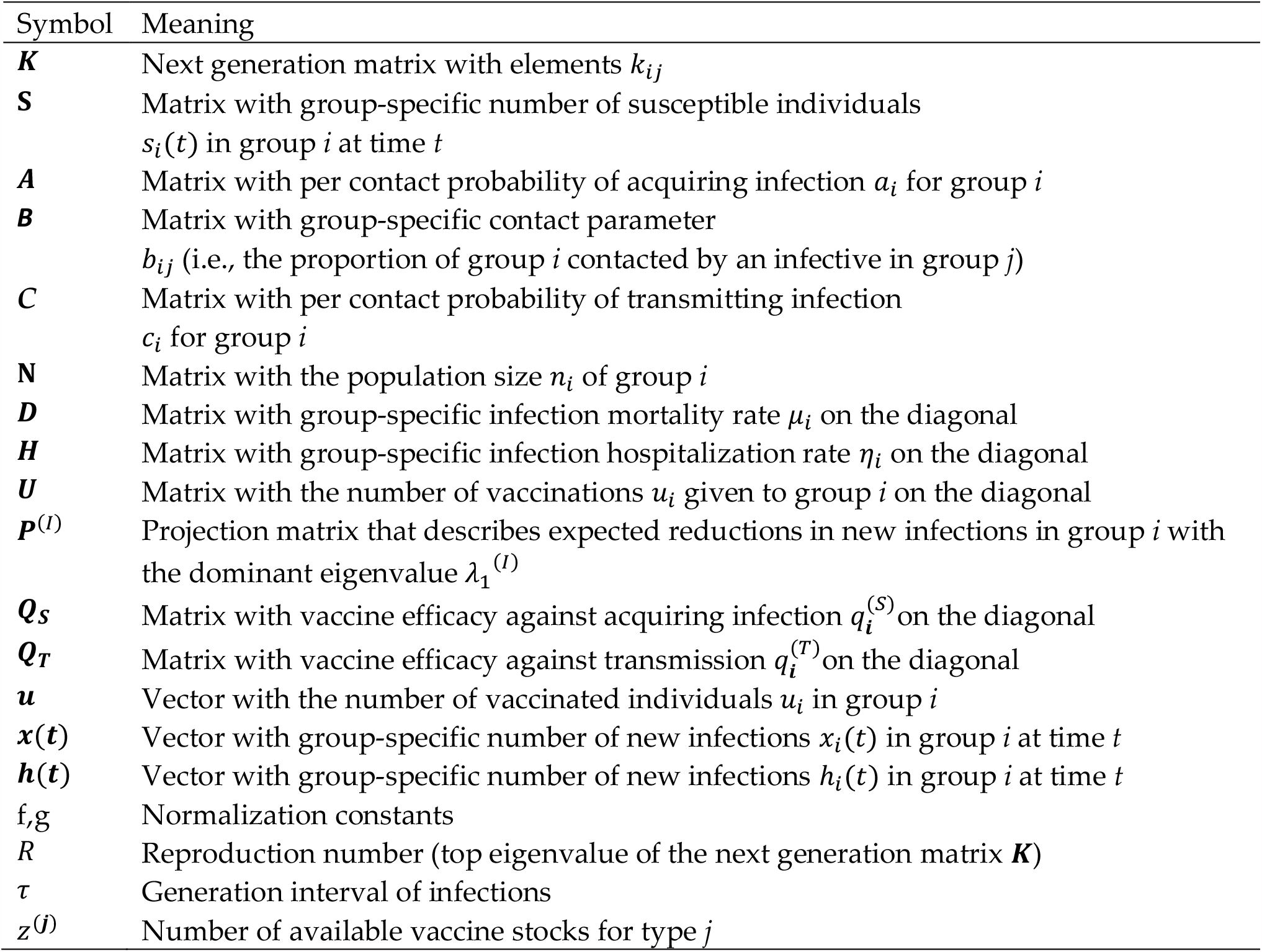
Notation and meaning of variables

**Table S2.**
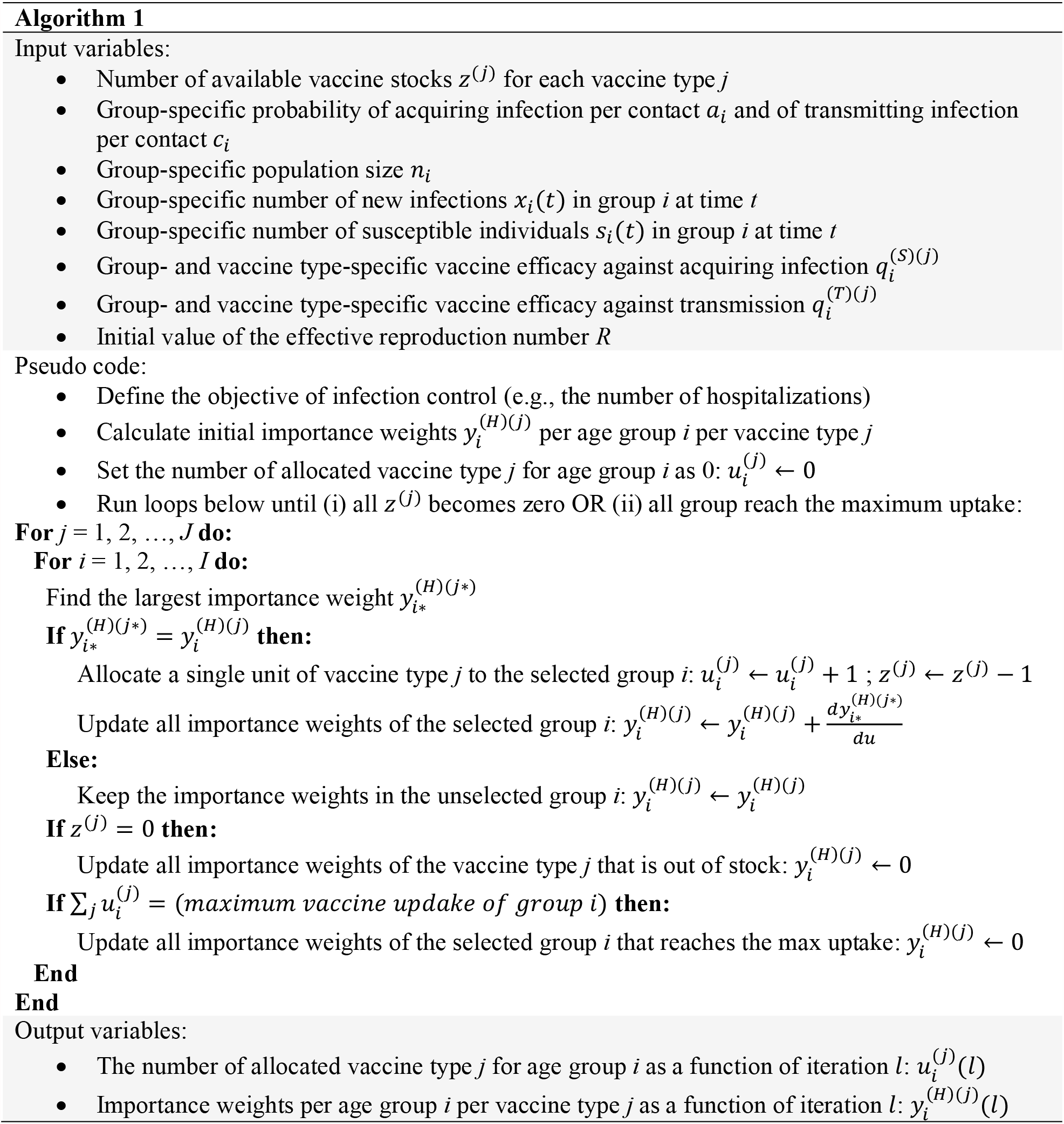
Pseudo code of the allocation algorithm

## Text S1. Mathematical details

### 1. Mathematical details

#### 1.1 Objective

The aim of following calculations is to formulate the expected impact of targeted vaccinations. We firstly present our approach to relate the expected changes in the next generation matrix ***K*** to the observed epidemiological data (i.e., the number of new infections per group). We then generalize the argument to quantify the expected impact in the number of hospitalizations and deaths.

The following analysis is known as “perturbation analysis” of a matrix in demography and population ecology (42,44), and we will refer to theorems and proofs introduced by those exiting literatures. For consistent notations, we follow Magnus and Neudecker (1988) (45); matrices are denoted by upper case bold symbols (e.g., **A**), and vectors are denoted by lower case bold symbols (**n**). Note that we define the derivatives of a matrix (or vector) as the matrix (or vector) of derivatives of the elements (e.g., 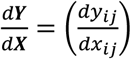 and 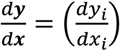). All notations and definitions of variables are shown in **Table-S2**.

#### 1.2 Next generation matrix

The host population is subdivided into *m* groups. The next generation matrix ***K*** gives the number of new infections in a successive generation, such that the number of new infections at time *t* + 1 after 1 generations of infections is ***x***(*t* + 1) = ***Kx***(*t*). For a large class of transmission models such as susceptible-infected-recovered model (SIR) model, the next generation matrix ***K*** can be written as

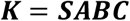

where matrices ***S, A, B***, and ***C*** have the following epidemiological interpretation: ***S*** is a matrix with group-specific number of susceptible individuals *s*_*i*_ (*t*) on the diagonal, ***A*** is a matrix with per contact probability of acquiring infection *a*_*i*_ on the diagonal, ***B*** is a contact matrix with elements *b*_*ij*_, and ***C*** is a matrix with group-specific per contact probability of transmitting infection *c*_*i*_ on the diagonal. Note that only ***S*** is time-dependent (and thus ***K*** is also time-dependent). For readability, when it is obvious from the context, we do not write the dependency on time. We require that at-risk contacts are reciprocal, and thus the matrix ***B*** is assumed to be symmetric. Thus, the next generation matrix ***K*** becomes a product of symmetric matrices and diagonalizable.

#### 1.3 Approximation by observed infections

By diagonalizing the next generation matrix ***K***, we have

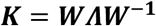

where ***Λ*** is a diagonal matrix that has eigenvalues *R, λ*_2_, *λ*_2_, …, *λ*_*m*_ as its elements and zeros elsewhere, where *R* is the dominant eigenvalue and is often referred to as the reproduction number. The matrix ***W*** has as columns the right eigenvectors ***w***_1_, ***w***_2_,…,***w***_*m*_. The matrix ***W***^−**1**^ is the inverse of the matrix ***W*** that has the left eigenvectors **v**_1_, **v**_2_,…,**v**_*m*_ as its rows. We require that an infector introduced in an arbitrary group reproduces a finite number of new infections in every group, and this condition ensures that the next generation matrix ***K*** is primitive. In such condition, the Perron-Frobenius Theorem guarantees that *R*, **w**_1_, and **v**_1_ are real and non-negative (25,42).

After *τ* generations of infections, the number of new infections at time *t* + *τ* is given by ***x***(*t* + *τ*) = ***K***^*τ*^***x***(*t*) = ***WΛ***^*τ*^***W***^−**1**^***x***(*t*). By using both right and left eigenvectors, we can rewrite the formula as

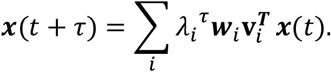

Note that ***x***(*t*) is a vector that has the number of new infections in age group *i* as its elements, denoted as *x*_*i*_(*t*) and that the dominant eigenvalue *λ*_1_ is the reproduction number *R*. If the dominant eigenvalue is strictly greater than other eigenvalues, the first term 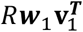 will eventually dominate other terms, and other all the terms will become negligible. This characteristic yields the approximated next generation matrix

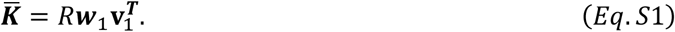

Now it is of interest to approximate the top right and left eigenvectors, **w**_1_ and **v**_1_, by observations. If the observation interval is long enough (typically longer than two generations of infections), we can safely approximate the top right eigenvector ***w***_1_ with the number of new infections ***x***(*t*) (28). Thus, we have

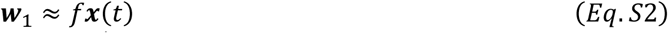

where *f* is the normalizing constant given by 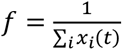. This result is also known as the strong ergodic theorem (see ref (46), p.86). Since the contact matrix **B** is symmetric and thus the next generation matrix ***K*** is a product of symmetric matrices, there exists a transformation matrix ***M*** that transposes ***K***, such that ***MKM***^−**1**^ = ***K***^***T***^. With this relationship, we can project the top left eigenvector **v**_1_ along the top right eigenvector **w**_1_, and subsequently

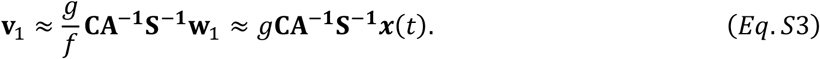

where *g* is the normalization constant given by 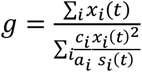. See detailed derivation in the section 3.4. of supporting info in (28).

#### 1.4 Sensitivity of the number of new infections to targeted vaccinations

##### 1.4.1 Changes in the number of new infections due to vaccinations

A decrease in ***x***(*t* + 1) is expressed as a result of changes in the next generation matrix ***K*** and in the number of infected individuals ***x***(*t*):

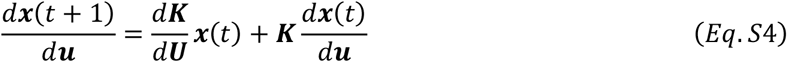

where 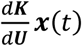 is the direct effect of vaccinating an individual and removing them from the susceptible population and 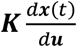 is the indirect effect of vaccinating a single individual by reducing onward infections.

##### 1.4.2 Perturbation in the next generation matrix K

We focus on the impact of vaccinations when vaccines are allocated to the group that can be immune or susceptible. The perturbation of next generation matrix 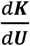 is expressed in terms of the change in the number of susceptible individuals 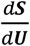 due to vaccinations, such that 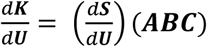. We denote the vaccine efficacy on susceptibility as ***Q***_***s***_, and the depletion of susceptible individual is written as

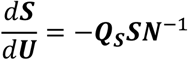

where ***N*** is a diagonal matrix that has elements of total population in each age group *n*_*i*_. Since 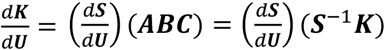, the perturbation of the next generation matrix is

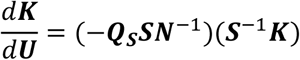

and thus

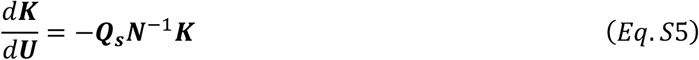

because ***S*** and ***N*** are diagonal matrices and thus commutative. The derivation is same as that of section 3.5. of supporting info in (28). Note that the vaccine efficacy of susceptibility here (i.e., ***Q***_***s***_) is defined as the probability of protecting infection per infectious contact (see next section 1.4.3 for another effect of vaccinations, which considers the prevention of transmission from an infectious individual).

##### 1.4.3 Perturbation in the number of infected individuals x(t)

If vaccines are allocated also to infected individuals ***x***(*t*) at time *t*, the change in the number of infected (infectious) individuals is

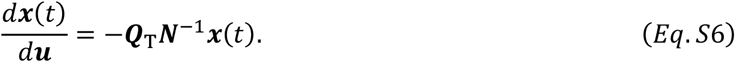

where ***Q***_T_ is the vaccine efficacy against the transmissibility.

##### 1.4.4 Importance weight of infection

By substituting Eq.S5 and Eq.S6 to Eq.S4, the decrease in the number of new infections after one generation is rewritten as

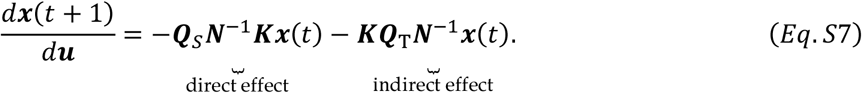

The interpretation of first term is the reduction in the number of new infections because susceptible individuals were depleted (i.e., direct effect), and that of second term is the effect preventing onward infections because infectious individuals are depleted (i.e., indirect effect).

Now we can relate this sensitivity 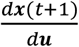 to observations. By approximating the next generation matrix by dominant right and left eigenvectors (i.e., 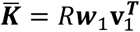), the above equation is rewritten as

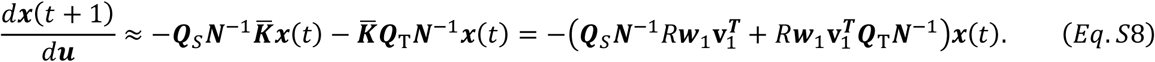

We set a projection matrix as 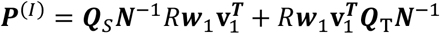 and its dominant eigenvalue as *λ*_1_^(*R*)^. When the vaccination is targeted to the group *i*, using Eq.S2 and S3, we obtain

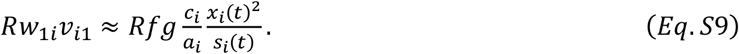

We use the same approximation method as section 1.3 for the projection matrix ***P***^(*I*)^ and its top right eigenvector 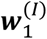. Given the sufficient length of observation intervals, we can safely approximate 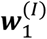 by the number of new infections ***x***(*t*) such that 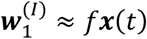 (see ref (46), p.86). Since 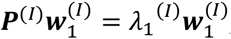, the contribution of age group *i* to the dominant eigenvalue *λ*_1_^(*I*)^ is:

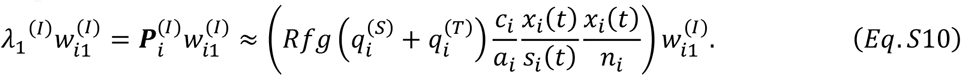

We can interpret the quantity 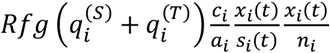 as the expected reduction in the number of new infections generated by a typical infected individual in group *i* after introducing a single unit of vaccine. Thus, we define this expected impact of a single vaccination in group *i* on the dominant eigenvalue *λ*_1_^(*I*)^ as the importance weight of infection:

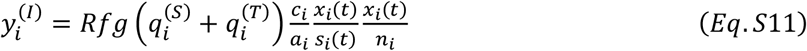

#### 1.5 Sensitivity of the number of hospitalizations to targeted vaccinations

##### 1.5.1 Changes in the number of hospitalizations due to vaccinations

The number of hospitalized individuals ***h***(*t*) at time *t* (i.e., *m* × 1 vector with elements *h*_1_, *h*_2_, …, *h*_*m*_) is defined as

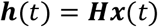

where ***H*** is a diagonal matrix with group-specific hospitalization rate *η*_1_, *η*_2_, …, *η*_*m*_. Suppose that we wish to predict the number of hospitalizations after one generation of infections. We can write the number of new hospitalizations as

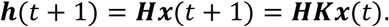

Over the observation interval from *t* to *t* + 1, we assume the group-specific probability of hospitalization is constant.

Here we look at the perturbation of the expected number of hospitalizations ***h***(*t* + 1) due to vaccinations. Since the infection hospitalization rate matrix ***H*** is constant, the perturbation in ***x***(*t* + 1) is of interest. Therefore, using Eq.S4, the decrease in the number of hospitalizations can be written as

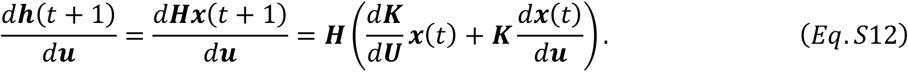

##### 1.5.2 Importance weight of hospitalization

Using Eq.S12 and the result of section 1.4.4 such as Eq.S8, the small change in the expected number of hospitalizations after one generation is now written as

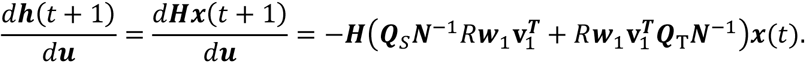

Again, we set a projection matrix as 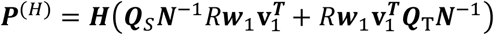 and its dominant eigenvalue as *λ*_1_^(*H*)^. When the vaccination is targeted to the group *i*, the relative change in the dominant eigenvalue *λ*_1_^(*H*)^ is

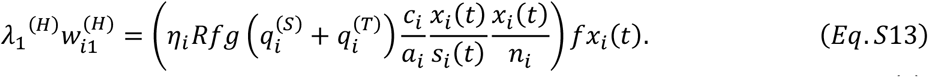

We define this expected impact of a single vaccination in group *i* on the dominant eigenvalue *λ*_1_^(*H*)^ as the importance weight of hospitalization:

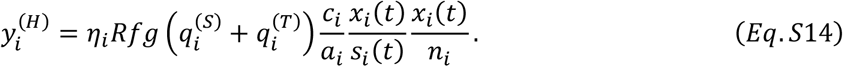

We can interpret this quantity 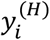 as the expected reduction in the number of new hospitalizations generated by a typical infected individual in group *i* after introducing a single unit of vaccine.

#### 1.6 Importance weights for other objectives

We can replace the matrix ***H*** (i.e., a diagonal matrix with the elements of infection hospitalization rates per group *i*) with different rate matrices for other objectives. In this study, we also aimed to test an allocation strategy to minimize the number of deaths. Thus, we introduced a diagonal matrix ***D*** with group-specific infection mortality rate *μ*_*i*_ on the diagonal, and the importance weight of death 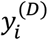 can be derived in the same manner as the section 1.5;

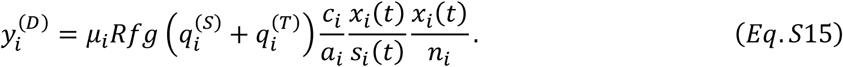

We can interpret this quantity 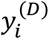 as the expected reduction in the number of new deaths generated by a typical infected individual in group *i* after introducing a single unit of vaccine.

#### 1.7 Changes in importance weights during the allocation of vaccines

Since importance weights are dependent on the number of allocated vaccines, we need to update them at each allocation step. We denote the changes in importance weights in group *i* per single allocation as 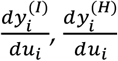, and 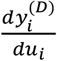 for each objective.

When vaccines are allocated, the eigenvector ***w***_1_ is perturbed, and its small change 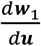 can be approximated using the Power iteration with the new matrix 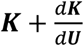 (see (47), p.331):

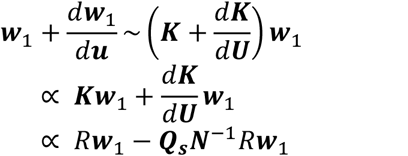

where the sign ∝ means “proportional to” and the sign ∼ means “approximately proportional to”. The same derivation has been introduced elsewhere (see section 3.6. of supporting info in (28)). If the allocation of vaccines is targeted to the group *i*, this results in the change in the *i* th element of the top right eigenvector, such as

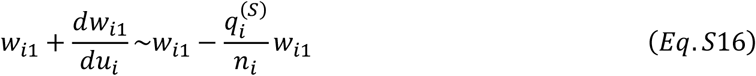

And all other elements remain unchanged (28). We use this equation to quantify the changes in importance weights after the allocation of a single unit of vaccines. By multiplying both sides of Eq.S16 by the factor 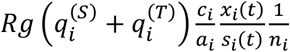 and using *w*_i1_ ≈ *fx*_*i*_ (*t*), from Eq.S2 and Eq.S11, we obtain

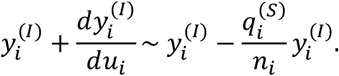

Subsequently, we can define the change in the importance weights of hospitalization and death. By multiplying both sides of Eq.S16 by factors 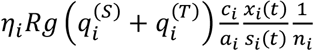 and 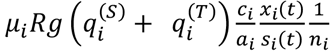 respectively, from Eq.S2 and Eq.S14-15, we obtain

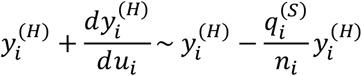

and

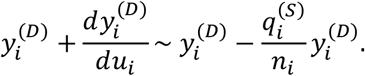

The perturbation due to vaccination in group *i* does not affect other groups.

## References

1. ECDC. COVID-19 situation update for the EU/EEA, as of week 26. European Centre for Disease Prevention and Control (ECDC). [cited 2021 Jul 9]. Available from: https://www.ecdc.europa.eu/en/cases-2019-ncov-eueea

2. WHO Coronavirus (COVID-19) Dashboard. [cited 2021 Jul 9]. Available from: https://covid19.who.int/

3. Kucharski AJ, Klepac P, Conlan AJK, Kissler SM, Tang ML, Fry H, et al. Effectiveness of isolation, testing, contact tracing, and physical distancing on reducing transmission of SARS-CoV-2 in different settings: a mathematical modelling study. Lancet Infect Dis. 2020 Oct;20(10):1151–60.

4. Ferretti L, Wymant C, Kendall M, Zhao L, Nurtay A, Abeler-Dörner L, et al. Quantifying SARS-CoV-2 transmission suggests epidemic control with digital contact tracing. Science. 2020 May 8;368(6491). Available from: http://dx.doi.org/10.1126/science.abb6936

5. Sandmann FG, Davies NG, Vassall A, Edmunds WJ, Jit M, Centre for the Mathematical Modelling of Infectious Diseases COVID-19 working group. The potential health and economic value of SARS-CoV-2 vaccination alongside physical distancing in the UK: a transmission model-based future scenario analysis and economic evaluation. Lancet Infect Dis. 2021 Mar 18; Available from: http://dx.doi.org/10.1016/S1473-3099(21)00079-7

6. Cutler DM, Summers LH. The COVID-19 Pandemic and the $16 Trillion Virus. JAMA. 2020 Oct 20;324(15):1495–6.

7. Shrotri M, Swinnen T, Kampmann B, Parker EPK. An interactive website tracking COVID-19 vaccine development. Lancet Glob Health. 2021 Mar 2; Available from: http://dx.doi.org/10.1016/S2214-109X(21)00043-7

8. Kluge H, McKee M. COVID-19 vaccines for the European region: an unprecedented challenge. Lancet. 2021 Mar 25; Available from: http://dx.doi.org/10.1016/S0140-6736(21)00709-1

9. Cowling BJ, Lim WW, Cobey S. Fractionation of COVID-19 vaccine doses could extend limited supplies and reduce mortality. Nat Med. 2021 Jul 5;1–2.

10. Mylius SD, Hagenaars TJ, Lugnér AK, Wallinga J. Optimal allocation of pandemic influenza vaccine depends on age, risk and timing. Vaccine. 2008 Jul 4;26(29–30):3742–9.

11. Medlock J, Galvani AP. Optimizing influenza vaccine distribution. Science. 2009 Sep 25;325(5948):1705– 8.

12. Bansal S, Pourbohloul B, Meyers LA. A comparative analysis of influenza vaccination programs. PLoS Med. 2006 Oct;3(10):e387.

13. Moore S, Hill EM, Tildesley MJ, Dyson L, Keeling MJ. Vaccination and non-pharmaceutical interventions for COVID-19: a mathematical modelling study. Lancet Infect Dis. 2021 Mar 18; Available from: https://www.sciencedirect.com/science/article/pii/S1473309921001432

14. Viana J, van Dorp CH, Nunes A, Gomes MC, van Boven M, Kretzschmar ME, et al. Controlling the pandemic during the SARS-CoV-2 vaccination rollout: a modeling study. medRxiv. medRxiv; 2021. Available from: http://medrxiv.org/lookup/doi/10.1101/2021.03.24.21254188

15. Giordano G, Colaneri M, Di Filippo A, Blanchini F, Bolzern P, De Nicolao G, et al. Modeling vaccination rollouts, SARS-CoV-2 variants and the requirement for non-pharmaceutical interventions in Italy. Nat Med. 2021 Apr 16; Available from: http://dx.doi.org/10.1038/s41591-021-01334-5

16. Bubar KM, Reinholt K, Kissler SM, Lipsitch M, Cobey S, Grad YH, et al. Model-informed COVID-19 vaccine prioritization strategies by age and serostatus. Science. 2021 Jan 21 [cited 2021 Jan 22]; Available from: https://science.sciencemag.org/content/early/2021/01/21/science.abe6959

17. Matrajt L, Eaton J, Leung T, Brown ER. Vaccine optimization for COVID-19: Who to vaccinate first? Science Advances. 2020 Feb 1;7(6):eabf1374.

18. Buckner JH, Chowell G, Springborn MR. Dynamic prioritization of COVID-19 vaccines when social distancing is limited for essential workers. Proc Natl Acad Sci U S A. 2021 Apr 20;118(16). Available from: http://dx.doi.org/10.1073/pnas.2025786118

19. Davies NG, Klepac P, Liu Y, Prem K, Jit M, CMMID COVID-19 working group, et al. Age-dependent effects in the transmission and control of COVID-19 epidemics. Nat Med. 2020 Aug;26(8):1205–11.

20. Zhang J, Litvinova M, Liang Y, Wang Y, Wang W, Zhao S, et al. Changes in contact patterns shape the dynamics of the COVID-19 outbreak in China. Science. 2020 Jun 26;368(6498):1481–6.

21. Salje H, Tran Kiem C, Lefrancq N, Courtejoie N, Bosetti P, Paireau J, et al. Estimating the burden of SARS-CoV-2 in France. Science. 2020 Jul 10;369(6500):208–11.

22. Walker PGT, Whittaker C, Watson OJ, Baguelin M, Winskill P, Hamlet A, et al. The impact of COVID-19 and strategies for mitigation and suppression in low-and middle-income countries. Science. 2020 Jul 24;369(6502):413–22.

23. Levin AT, Hanage WP, Owusu-Boaitey N, Cochran KB, Walsh SP, Meyerowitz-Katz G. Assessing the age specificity of infection fatality rates for COVID-19: systematic review, meta-analysis, and public policy implications. Eur J Epidemiol. 2020 Dec 8; Available from: http://dx.doi.org/10.1007/s10654-020-00698-1

24. O’Driscoll M, Ribeiro Dos Santos G, Wang L, Cummings DAT, Azman AS, Paireau J, et al. Age-specific mortality and immunity patterns of SARS-CoV-2. Nature. 2020 Nov 2; Available from: http://dx.doi.org/10.1038/s41586-020-2918-0

25. Diekmann O, Heesterbeek JAP. Mathematical Epidemiology of Infectious Diseases: Model Building, Analysis and Interpretation. John Wiley & Sons; 2000. 303 p.

26. Diekmann O, Heesterbeek JAP, Roberts MG. The construction of next-generation matrices for compartmental epidemic models. J R Soc Interface. 2010 Jun 6;7(47):873–85.

27. van den Driessche P. Reproduction numbers of infectious disease models. Infect Dis Model. 2017 Aug;2(3):288–303.

28. Wallinga J, van Boven M, Lipsitch M. Optimizing infectious disease interventions during an emerging epidemic. Proc Natl Acad Sci U S A. 2010 Jan 12;107(2):923–8.

29. Government of the Netherlands. Approach to corona vaccination in the Netherlands. 2021 [cited 2021 Jul 9]. Available from: https://www.rijksoverheid.nl/onderwerpen/coronavirus-vaccinatie/aanpak-coronavaccinatie/kinderen-en-jongeren

30. Cormen TH, Leiserson CE, Rivest RL, Stein C. Introduction To Algorithms. MIT Press; 2001. 1180 p.

31. Feehan DM, Mahmud AS. Quantifying population contact patterns in the United States during the COVID-19 pandemic. Nat Commun. 2021 Feb 9;12(1):1–9.

32. Backer JA, Mollema L, Vos ER, Klinkenberg D, van der Klis FR, de Melker HE, et al. Impact of physical distancing measures against COVID-19 on contacts and mixing patterns: repeated cross-sectional surveys, the Netherlands, 2016-17, April 2020 and June 2020. Euro Surveill. 2021 Feb;26(8). Available from: http://dx.doi.org/10.2807/1560-7917.ES.2021.26.8.2000994

33. Roser M, Ritchie H, Ortiz-Ospina E, Hasell J. Coronavirus pandemic (COVID-19). OurWorldInData.org. 2021 [cited 2021 Jul 12]. Available from: https://ourworldindata.org/coronavirus

34. Russell TW, Golding N, Hellewell J, Abbott S, Wright L, Pearson CAB, et al. Reconstructing the early global dynamics of under-ascertained COVID-19 cases and infections. BMC Med. 2020 Oct 22;18(1):332.

35. Omori R, Mizumoto K, Nishiura H. Ascertainment rate of novel coronavirus disease (COVID-19) in Japan. Int J Infect Dis. 2020 Jul;96:673–5.

36. Wallinga J, Teunis P, Kretzschmar M. Using data on social contacts to estimate age-specific transmission parameters for respiratory-spread infectious agents. Am J Epidemiol. 2006 Nov 15;164(10):936– 44.

37. Vos ERA, van Boven M, den Hartog G, Backer JA, Klinkenberg D, van Hagen CCE, et al. Associations between measures of social distancing and SARS-CoV-2 seropositivity: a nationwide population-based study in the Netherlands. medRxiv. medRxiv; 2021. Available from: http://medrxiv.org/lookup/doi/10.1101/2021.02.10.21251477

38. Polack FP, Thomas SJ, Kitchin N, Absalon J, Gurtman A, Lockhart S, et al. Safety and Efficacy of the BNT162b2 mRNA Covid-19 Vaccine. N Engl J Med. 2020 Dec 10; Available from: https://doi.org/10.1056/NEJMoa2034577

39. Baden LR, El Sahly HM, Essink B, Kotloff K, Frey S, Novak R, et al. Efficacy and Safety of the mRNA-1273 SARS-CoV-2 Vaccine. N Engl J Med. 2020 Dec 30; Available from: http://dx.doi.org/10.1056/NEJMoa2035389

40. Voysey M, Clemens SAC, Madhi SA, Weckx LY, Folegatti PM, Aley PK, et al. Safety and efficacy of the ChAdOx1 nCoV-19 vaccine (AZD1222) against SARS-CoV-2: an interim analysis of four randomised controlled trials in Brazil, South Africa, and the UK. Lancet. 2021 Jan 9;397(10269):99–111.

41. Oliver SE, Gargano JW, Scobie H, Wallace M, Hadler SC, Leung J, et al. The Advisory Committee on Immunization Practices’ Interim Recommendation for Use of Janssen COVID-19 Vaccine - United States, February 2021. MMWR Morb Mortal Wkly Rep. 2021 Mar 5;70(9):329–32.

42. Caswell H. Sensitivity Analysis: Matrix Methods in Demography and Ecology. Springer, Cham; 2019.

43. Blower SM, Dowlatabadi H. Sensitivity and Uncertainty Analysis of Complex Models of Disease Transmission: An HIV Model, as an Example. Int Stat Rev. 1994;62(2):229–43.

44. Inaba H. Age-Structured Population Dynamics in Demography and Epidemiology. Springer; 2017. 555 p.

45. Magnus JR, Neudecker H. Matrix Differential Calculus with Applications in Statistics and Econometrics. Wiley; 1988. 393 p.

46. Caswell H. Matrix population models. Vol. 1. Sinauer Sunderland, MA, USA; 2000.

47. Golub GH, Van Loan CF. Matrix computations (3rd ed.). USA: Johns Hopkins University Press; 1996.

